# Validation and Convergent Validity of the Boston Cognitive Assessment (BOCA) in an Italian Population: A Comparative Study with the Montreal Cognitive Assessment (MoCA) in Alzheimer’s Disease Spectrum

**DOI:** 10.1101/2024.05.05.24306896

**Authors:** Alessandro Padovani, Salvatore Caratozzolo, Alice Galli, Luca Crosani, Silvio Zampini, Maura Cosseddu, Rosanna Turrone, Andrea Zancanaro, Bianca Gumina, Barbara Vicini-Chilovi, Alberto Benussi, Andrey Vyshedskiy, Andrea Pilotto

**Author notes:** Corresponding author: Salvatore Caratozzolo MD, Neurology Unit, University of Brescia, P. le Spedali Civili 1, 25123, Brescia, Italy. All authors contributed to the study conception and design. Material preparation, data collection and analysis were performed by [full name], [full name] and [full name]. The first draft of the manuscript was written by [full name] and all authors commented on previous versions of the manuscript. All authors read and approved the final manuscript. All authors certify that they have no affiliations with or involvement in any organization or entity with any financial interest or non-financial interest in the subject matter or materials discussed in this manuscript.

## Abstract

**Background:** The Boston Cognitive Assessment (BOCA) is a self-administered online test developed for cognitive screening and longitudinal monitoring of brain health in an aging population. The study aimed to validate BOCA in an Italian population and to investigate the convergent validity with the Montreal Cognitive Assessment (MOCA) in normal controls and subjects within the Alzheimer Disease spectrum.

**Methods:** BOCA was administered to 150 participants, including cognitively healthy controls (HC, n=50), patients with mild cognitive impairment (MCI, n=50), and dementia (DEM, n=50). The BOCA reliability was assessed using i) Spearman’s correlation analysis between subscales; ii) Cronbach’s alpha calculation, and iii) Principal Component Analysis. Repeated-measures ANOVA was employed to assess the impact of the sequence of test administrations between the groups. BOCA performance between HS, MCI and DEM were compared using Kruskall Wallis test. Furthermore, a comparison was conducted between MCI patients who tested positive for amyloid and those who tested negative, utilizing Mann Whitney’s U-test.

**Results:** Test scores were significantly different between patients and controls (p<0.001) suggesting good discriminative ability. The Cronbach’s alpha was 0.82 indicating a good internal consistency of the BOCA subscales and strong-to-moderate Spearman’s correlation coefficients between them. BOCA demonstrated strong correlation with Montreal Cognitive Assessment (MoCA) (rho=0.790, p<0.001).

**Conclusions:** The Italian version of the BOCA test exhibited validity, feasibility, and accurate discrimination.

## BACKGROUND

The assessment of cognitive decline and the prediction of dementia risk remain crucial aspects in addressing the challenges posed by an aging population. The expanding population of elderly individuals proficient in smartphone and digital technology offers significant opportunities for utilizing digital cognitive assessments in unsupervised settings (Öhman et al., 2021) [1]. Over the past decade, there has been a notable surge in smartphone-based applications for cognitive assessment (Koo et al., 2019) [2]. The advantages of unsupervised computerized cognitive tests are manyfold, as they provide standardized administration, minimizing subjective influences from examiners and ensuring consistent evaluation across participants; they reduce performance anxiety and the so-called “white-coat effect” observed in medical settings and thus potentially leading to more accurate results. They have the potential to reduce the time required from examiners and enable remote administration, thereby reducing the necessity for in-person appointments and related travel expenses. This remote administration makes cognitive assessments more cost-effective and accessible, even in primary care settings. Furthermore, longitudinal monitoring of cognitive health can help clinicians assess if an underlying condition is causing cognitive decline and guide timely therapeutic interventions (Kim J, et al. 2017) [3]. In addition, follow-up monitoring is essential for testing novel interventions designed to reduce or reverse cognitive aging (Foster et al. 2019) [4]. Computerized tests automatically record and store assessment data, thus reducing the likelihood of errors associated with manual data entry and facilitating easy access to longitudinal performance data for tracking changes over time; (Samaroo et al., 2019, Öhman et al., 2021, Zygouris et al. 2015, Ruoano et al. 2015) [1,5,6]. Recently, a self-administered 10-min-digital tool called Boston Cognitive Assessment (BOCA) has been developed (Vyshedskiy et al. 2022) [7] demonstrating good internal consistency, adequate content validity, and strong test-retest reliability. BOCA demonstrated strong correlation with Montreal Cognitive Assessment (MoCA) and potentially represents a valid tool for early detection and monitoring of cognitive deficits, particularly in individuals at risk of cognitive impairments and dementia (Vyshedskiy et al., 2022) [7].

In this study, we aim at evaluating the Italian version of BOCA and test its validity and feasibility in a large group of participants including healthy elderly, individuals with mild cognitive impairment and patients with mild to moderate dementia.

## METHODS

### Participants

The study included individuals with cognitive deficits consecutively recruited by the Memory Clinic of the University of Brescia and a group of controls matched for age, sex, and educational levels recruited among the patients’ caregivers. The study was approved by the Local Ethic Committee (NP 1471, DMA, Brescia, approved in its last version on April 19th, 2022). Each participant gave their informed consent prior to their inclusion. Cognitive impairment was defined by subjective cognitive complaints over a period of at least 6 months reported by the patient or caregivers. Among patients with cognitive deficits, individuals were assigned to the mild cognitive impairment (MCI) (Petersen & Negash, 2008) [8] group if their Clinical Dementia Rating scale (CDR) (Hughes et al., 1982) [9] was less than one; patients with CDR ≥ 1 were assigned into the Dementia group. Patients with MCI and dementia were further classified as amyloid-positive or -negative, according to the positivity of CSF pattern or amyloid-PET (Padovani et al., 2022) [10]. The functional abilities of participants were characterized using the basic activities of daily living (BADL) and the instrumental activities of daily living (IADL) (Lawton & Brody, 1969) [11].

Participants completed both BOCA and MoCA tests in a random order. The order-effect was assessed using a mixed effect model. All patients were unsupervised during the BOCA test administration, which was conducted in a quiet room at the hospital. Only a subset of cognitively healthy subjects completed the BOCA test at home.

### Statistical analyses

The two-sample t-test and chi-squared test were used to assess differences in demographic variables for continuous and categorical variables, respectively. The Shapiro-Wilk test was used to assess normal distribution. All subtest scores did not meet the normality assumption with p<0.05. To assess the correlation between BOCA and MoCA test scores, Spearman’s correlation was used with p<0.05 as the statistical threshold. Linear regression was then calculated separately for patients and controls to assess the relationship between total BOCA score (independent variable) and participant age or education (dependent variables). BOCA reliability analysis was performed in several steps. First, the correlation matrix between the BOCA subscales was generated using Spearman’s correlation. Correlation strengths were determined as follows: 0.1-0.3 indicated a weak association, 0.31-0.50 a moderate association and 0.51-1.0 a strong association. Secondly, Cronbach’s alpha was obtained to determine the internal consistency of the BOCA test. Specifically, Cronbach’s alpha was derived considering all eight subscales. Then, one subscale at a time was removed and Cronbach’s alpha was recalculated for the remaining 7 subscales. All subscale scores were standardized for the purpose of this test. Thirdly, principal component analysis (PCA) was used to determine the number of components explaining the variance of the BOCA subscales. The Kaiser-Meyer-Olkin measure of sampling adequacy was used to test the assumptions. Mixed models were then used to simultaneously account for differences between diagnosis (between-subjects factor) and order of test administration (within-subjects). Differences in MoCA and BOCA scores between patients and controls were assessed using the Kruskal-Wallis non-parametric test. Post-hoc comparisons to assess differences between patients with MCI and dementia were performed using the Mann-Whitney U test. Moreover, the same statistical design was applied to assess differences between MCI patients who resulted amyloid positive vs. negative. All statistical analyses were performed using R version 4.3.1 and Jasp version 18.1. The statistical threshold was set at p<0.05 for all tests.

### Data security

As previously described (Vyshedskiy et al. 2022) [7], the data in transit is encrypted using SSL. SSL stands for Secure Sockets Layer, a security protocol that creates an encrypted link between a web server and a web browser. SSL certificates secure online transactions and keep all information private and secure. The test data are stored in the secure cloud database in a reputable cloud provider.

## Results

### Baseline demographics and clinical features of the sample

BOCA and MoCA tests were administered to a total of 150 participants, namely 50 patients with MCI, 50 patients with dementia and 50 cognitively healthy controls, matched for age, sex, and educational levels (Table 1). Further, 33/50 (66%) of MCI and 49/50 (98%) of patients with dementia were classified as amyloid positive according to CSF amyloid-β 42 or amyloid-PET. MCI and Dementia patients differed from HC in all cognitive and functional measures (table 1)

**Table 1.** Demographics and clinical characteristics in patients and controls.

|  | Healthy Controls<br>(n = 50) | MCI<br>(n = 50) | Dementia<br>(n = 50) | p-value |
| --- | --- | --- | --- | --- |
| Age (mean yrs) | 70.8 (11.9) | 72.5 (6.9) | 72.1 (9.6) | 0.439 |
| Education (mean yrs) | 9.9 (4.1) | 8.9 (3.4) | 8.6 (3.2) | 0.082 |
| Female (%) | 20 (38.5) | 26 (52.0) | 33 (61.1) | 0.116 |
|  |  | Clinical features |  |  |
| CDR global | 0.0 (0.0) | 0.5 (0.0) | 1.5 (0.5) | <0.001 |
| IADL (lost) | 0.0 (0.0) | 1.1 (0.2) | 1.8 (0.4) | <0.001 |
| BADL (lost) | 0.0 (0.0) | 0.2 (0.4) | 1.4 (0.6) | <0.001 |
| BOCA Total Score | 25.4 (3.5) | 23.5 (2.2) | 17.6 (2.9) | <0.001 |
| MOCA Total Score | 27.8 (1.6) | 24.6 (3.4) | 16.1 (3.3) | <0.001 |
| Amyloid positive (%) | 0 (0) | 33 (66) | 49 (94) | <0.001 |

Data are expressed as mean (SD). Abbreviations: CDR, Clinical Dementia Rating scale; IADL, Instrumental Activities of Daily Living; BADL, Basic Activities of Daily Living. IADL and BADL scores are expressed as skills lost as respect to the total. Amyloid-positivity was established based on CSF amyloid-β 42 < 650 or the presence of amyloid-plaques assessed with <sup>11</sup>C-PiB PET.

**Table 2.** Internal consistency of the eight BOCA subscales.

|  | Cronbach's alpha (if item deleted) |
| --- | --- |
| Memory/ Immediate Recall | 0.807 |
| Language | 0.810 |
| Mental Rotation | 0.806 |
| Clock Test | 0.817 |
| Attention | 0.796 |
| Mental Math | 0.819 |
| Orientation | 0.818 |
| Memory/ Delayed Recall | 0.807 |

Linear regression models revealed a significant relationship between BOCA total scores and age (β=-0.377, p=0.005) or education (β= 0.322, p=0.020) only in the control group. The Spearman’s Correlation revealed a positive and strong association between MoCA and BOCA total scores (rho=0.790, p<0.001, CI 95% [0.723, 0.837]) considering the whole cohort (figure 1).

**Figure 1.**
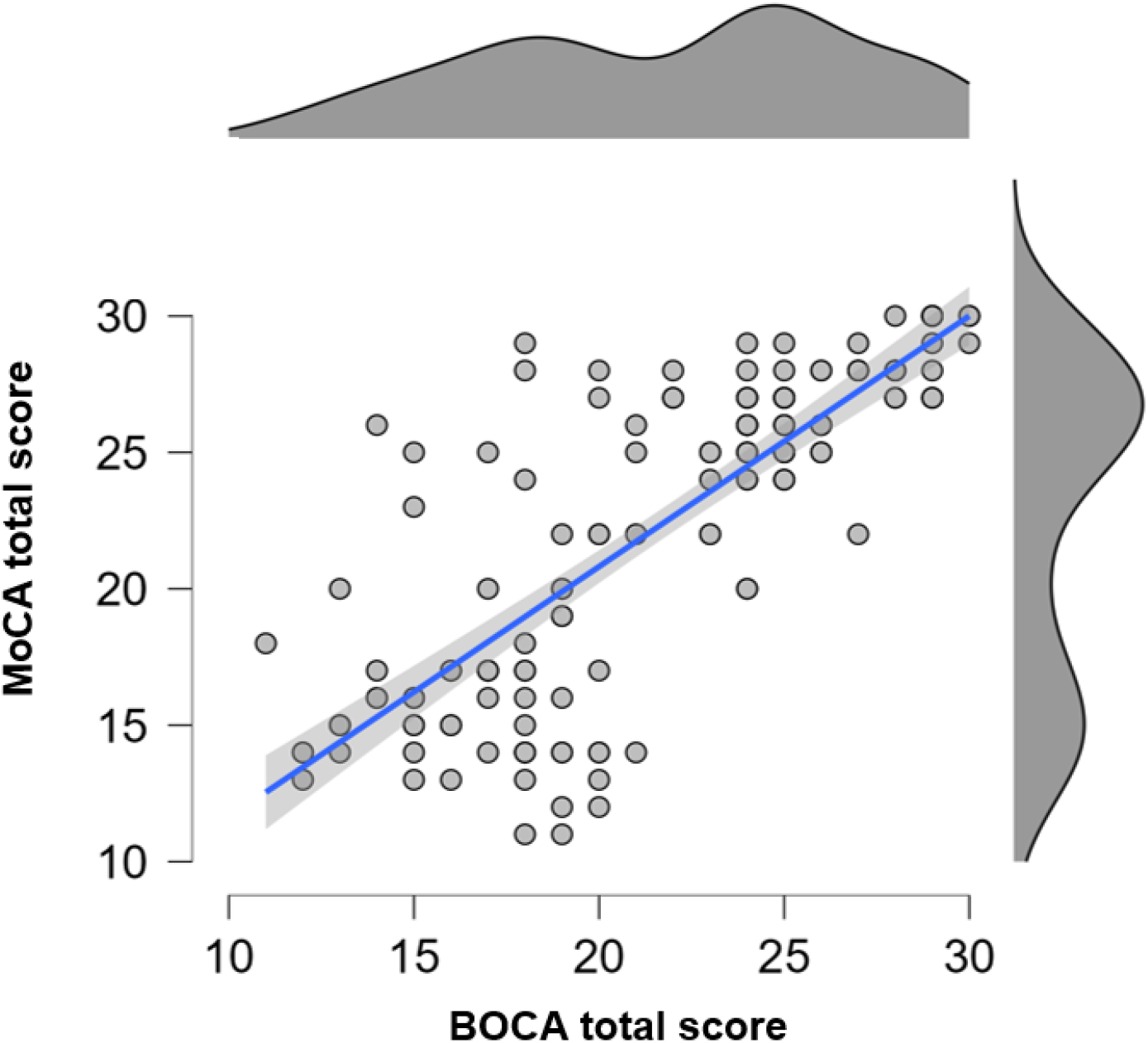
Correlation between MoCA and BOCA tests. Scatterplot representing the Spearman’s correlation between the two tests in the whole sample.

### BOCA reliability analysis

The correlation matrix for BOCA subscales was obtained via Spearman’s correlation method. The strongest correlation was between Orientation and Delayed Memory subscales (rho=0.51, p<0.001). Moderate associations were observed between Mental Math and Attention subscales (rho=0.451, p<0.001), Immediate Memory and Attention subscales (rho=0.438, p<0.001), Attention and Mental Rotation subscales (rho=0.435, p<0.001), and Mental Rotation and Language subscales (rho=0.424, p<0.001). Non-significant correlations were observed between Orientation and Mental Math subscales, and between Immediate Memory and Clock Test (see Figure 2).

**Figure 2.**
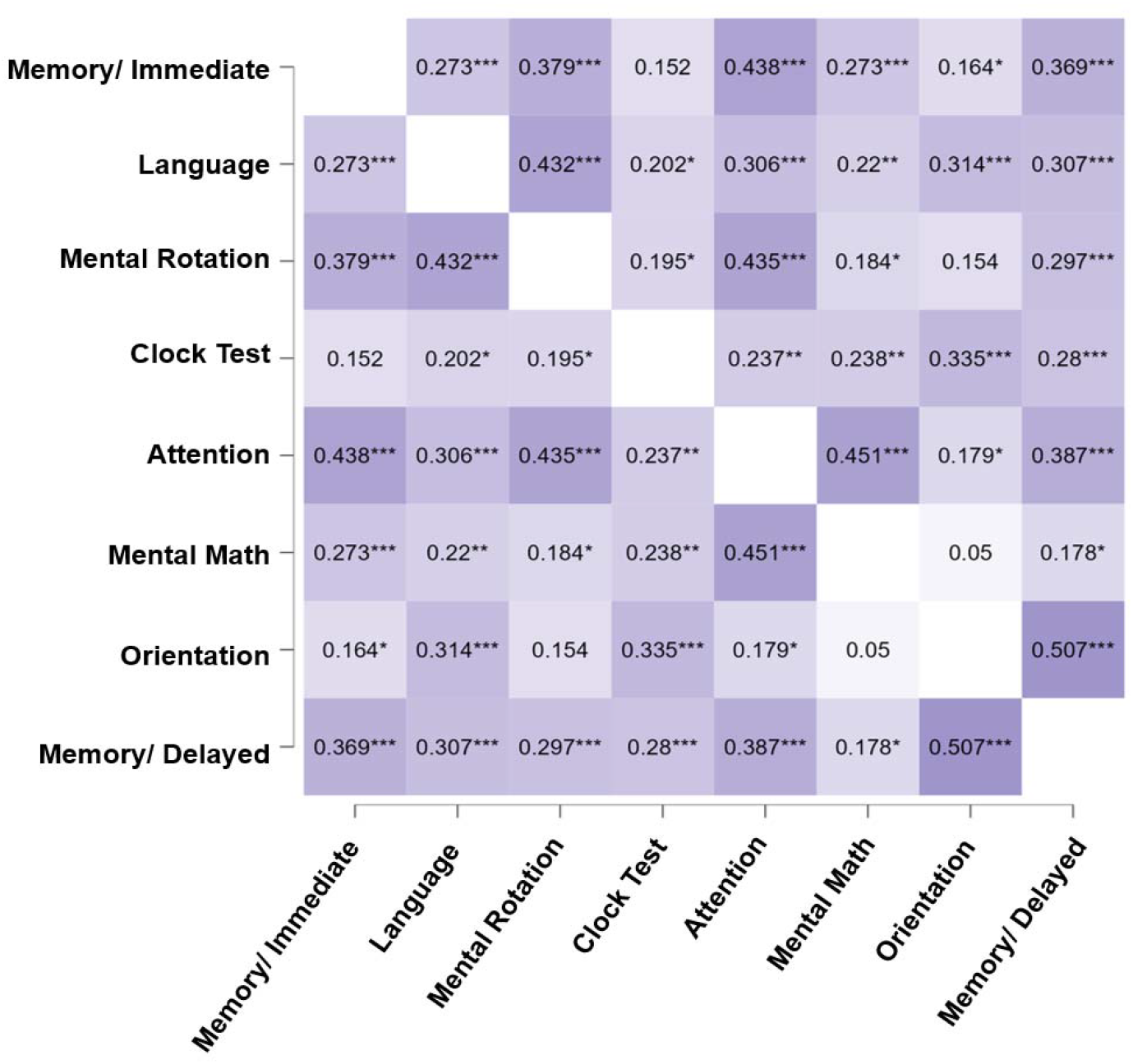
BOCA subscales correlation matrix. Heatmap representing significant Spearman correlations between BOCA subscales. Darker color represents a stronger relationship between variables. * = p<0.05; **= p<0.01; ***= p<0.001.

Internal consistency of the eight BOCA subscales was assessed using Cronbach’s alpha. Results indicated a good internal consistency (alpha= 0.822, CI 95% [0.775, 0.861]). Then, one subscale at time was removed, to re-calculate the Cronbach’s alpha for the remaining seven subscales. All subscales were standardized (i.e., z-scores) for this analysis. The resulting Cronbach’s alpha demonstrated high (>0.796) internal consistency (Table 5).

The Kaiser-Meyer-Olkin measure of sampling adequacy indicated that the strength of the relationship between the subscales was high (KMO=0.691) and the principal component analysis was possible. The eight subscales entered the model, and one component was retained using the eigenvalue > 1 criterion (eigenvalue=2.89), explaining the 37% of variance. All subscales significantly correlated with the selected component (p<0.05).

### Order effect assessment

Mixed models revealed no significant effect of the order of tests administration, also considering diagnosis as a between-subjects factor (BOCA total score: F=0.042, p=0.355; MoCA total score: F=0.259, p=0.772).

### Differences in the test scores between patients and controls

All participants completed MoCA and BOCA tests. The average MoCA score was 27.8 (SD=1.6) in controls, 24.6 (SD=3.4) in MCI and 16.1 (3.3) in patients with dementia. The MoCA total score was statistically different between the three considered groups (p<0.001). Visuo-spatial, Calculation, Language Fluency, Abstraction, and Delayed Recall subscales were significantly lower in MCI patients as compared to controls. Each subscale significantly differentiated between patients with dementia and controls (Table 3).

**Table 3.**
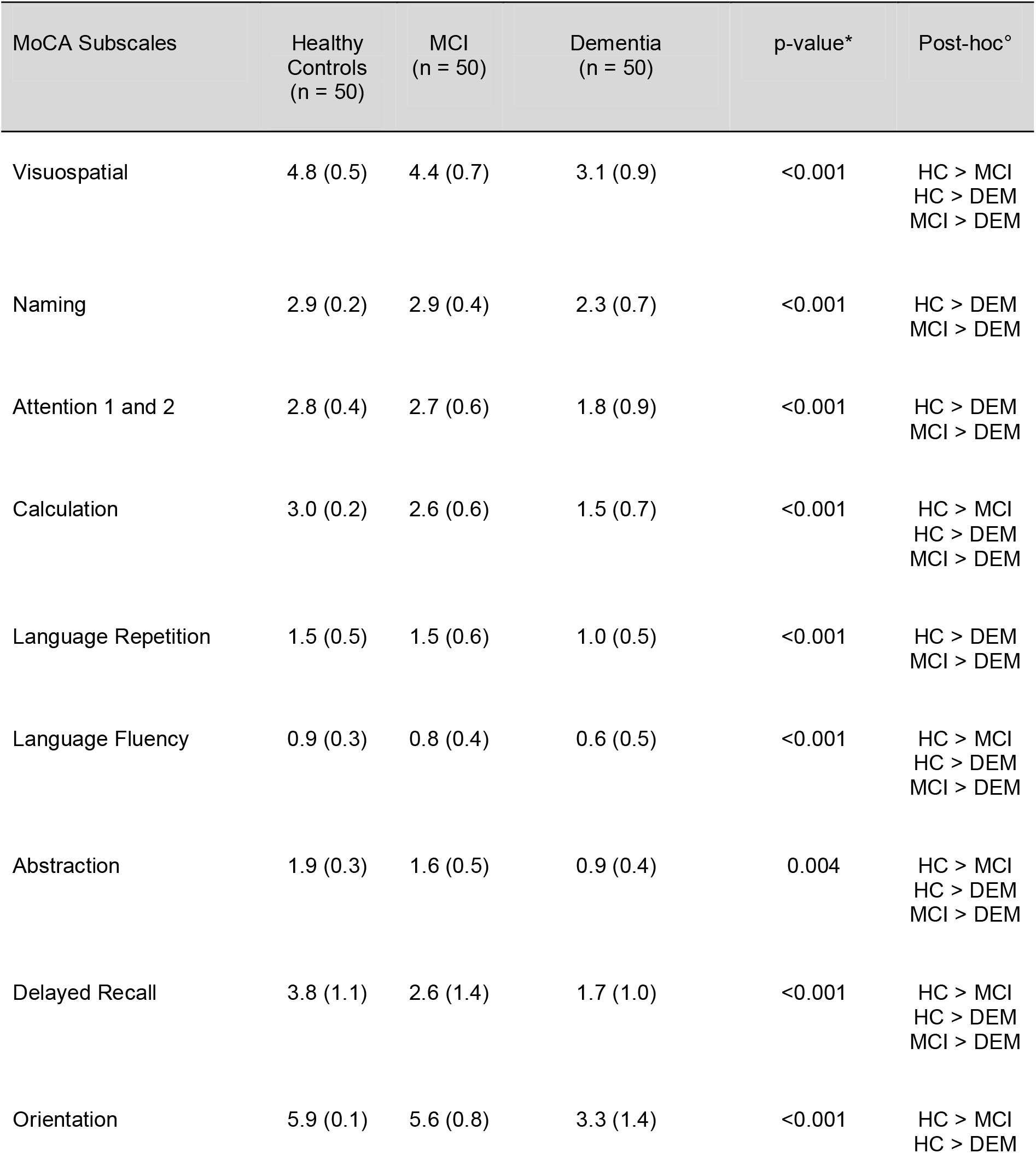
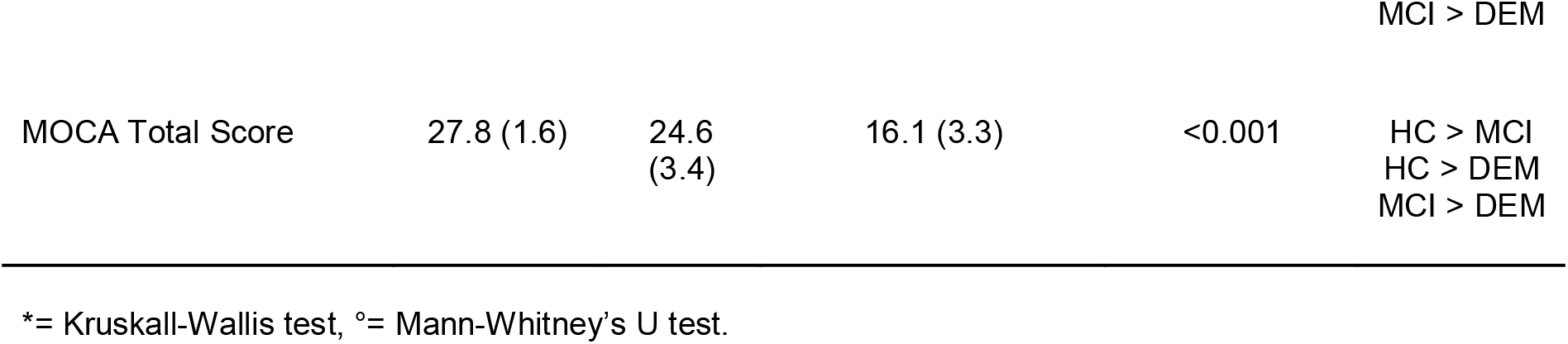
MoCA performance in patients and controls.

The average BOCA was 25.4 (SD= 3.5) for controls, 23.5 (SD=2.2) for MCI patients, and 17.6 (SD=2.9) for patients with dementia. The BOCA total score was significantly different between the three groups of individuals (p<0.001). Immediate and Delayed Memory, Attention, and Mental Math subscales significantly distinguished MCI patients from controls. All BOCA subscale scores were also significantly different between patients with dementia and controls (Table 4).

**Table 4.**
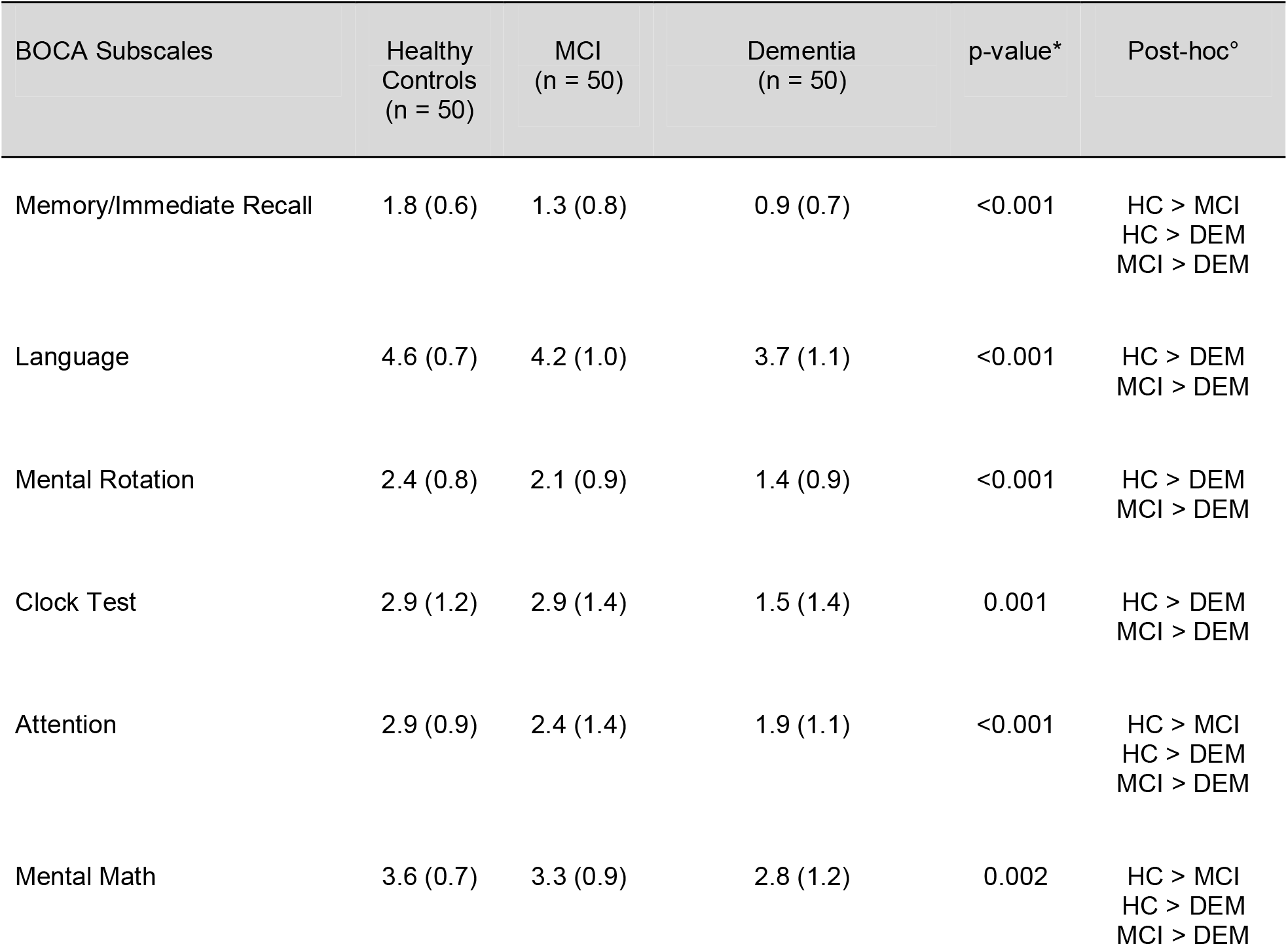
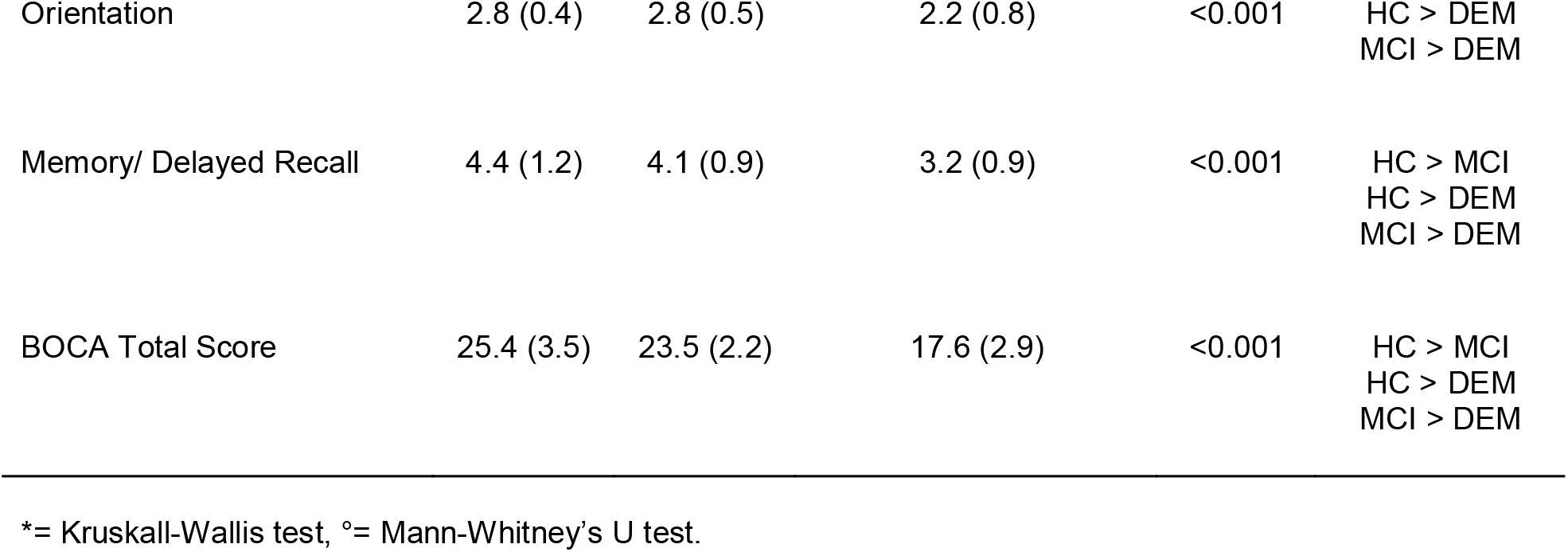
BOCA performance in patients and controls.

### Differences between amyloid positive and negative MCI patients

MCI amyloid-positive (MCI-Amy+, n=33) and MCI amyloid-negative (MCI-Amy-, n=17) were compared as for BOCA and MoCA. These two subgroups were comparable for age, sex, and education level. As for the MoCA test, only Abstraction subscale was significantly more impaired in MCI-Amy+ than MCI-Amy-(p=0.014). The MoCA total score in MCI-Amy+ was not statistically different from in MCI-Amy-(p=0.315).

Memory Immediate Recall (p=0.004), Memory Delayed recall (p=0.023), and Mental Rotation (p=0.045) BOCA subscales were significantly more impaired in MCI-Amy+ than MCI-Amy-. Consistently, the BOCA total score was lower in MCI-Amy+ than MCI-Amy-(p=0.048).

## Discussion

The study focused on the validation and feasibility of the Boston Cognitive Assessment (BOCA) in the Italian population, in a large set of subjects with normal cognition, MCI and mild to moderate dementia. The results of the study indicated a strong correlation between BOCA and MoCA total scores, showcasing good concurrent validity. The internal consistency of BOCA subscales was demonstrated to be high, with significant correlations observed between them. Both MoCA and BOCA tests were effective in differentiating between groups of healthy individuals, mild cognitive impairment, and dementia. These data agree with previous validation studies (Vyshedskiy et al 2022, Gold et al. 2022) [7,12] and support the usefulness of BOCA as a screening tool for the detection of cognitive impairment.

Over the last decade, there has been a remarkable increase in the development of digitalized tools for cognitive assessment (Öhman et al. 2021, Padovani 2023, Perin et al. 202, Kourtis et al. 2019, Zygouris et al. 2015) [1,5, 13-15]. Unsupervised computerized cognitive tests offer standardized administration, minimizing the subjective influences of examiners and ensuring consistent evaluation across participants. Still, the application of such digital testing is limited in clinical practice due to lack of feasibility studies testing their construct validity, applicability, and discriminative performances in real-world populations (Cubillos et al. 2023) [16]. In this study, 150 participants (50 healthy controls, 50 participants with MCI, and 50 with dementia) were included and underwent MoCA and unsupervised BOCA testing in a randomized order. The general applicability of the test was high, with no patients refusing the test or not being able to complete it by themself. BOCA and MoCA total scores demonstrated high correlation when considering the entire cohort. The internal consistency of BOCA subscales was high as demonstrated by the overall Cronbach’s alpha, even when each subscale was removed one at a time. Moreover, the principal component analyses revealed that BOCA subscales collectively represent a single underlying factor that explains a significant portion of the variability in the data. The results are very similar to the original validation performed by Vyshedskiy and coauthors (2022) [7] in the English version of BOCA and showed high applicability even in patients with MCI and dementia. Relative to the control group, a significant relationship between BOCA total scores and both age and education was found, as revealed by linear regression models. This finding is partially in contrast with previous literature in which BOCA was not found to be significantly associated with demographic characteristics (Gold D, et al. 2021) [12], whereas more recent validation studies showed an effect of education in the English version of BOCA (Ferguson et al. 2022) [17].

Additionally, three BOCA subscales (Memory Immediate Recall, Memory Delayed recall, and Mental Rotation) were notably lower in amyloid-positive patients compared to amyloid-negative individuals. While this alone may not be sufficient to determine BOCA’s ability to identify MCI due to Alzheimer’s Disorder (AD), it sheds light on the potential of the Boston Cognitive Assessment (BOCA) to discern subtle cognitive differences potentially associated with AD and need to be verified in longitudinal studies (Williams et al. 2020) [18].

Several limitations of the study should be acknowledged. Firstly, the sample size was fair but still limited, which may restrict the generalizability of our findings. Although we matched participants based on age, sex, and education level to mitigate potential confounding variables, a larger sample size would enhance the robustness of our results and allow for more comprehensive subgroup analyses. Another limitation is the lack of a control group with completely remote home-testing. It is important to consider the potential for uncontrolled bias in home-based settings, which may differ from the application in a quiet hospital room. Future research could compare BOCA scores obtained in controlled clinical settings with those obtained in at-home environments to assess potential discrepancies and identify strategies to minimize distractions during self-administration.

Additionally, it should be noted that our study primarily focused on patients with mild cognitive impairment (MCI) and dementia. Therefore, the generalizability of our findings to other patient populations may be limited. To address this limitation, future research should investigate BOCA scores in diverse patient groups, such as those with stroke, traumatic brain injury, aphasia, and other neurological disorders. Comparing BOCA scores across different patient populations would provide valuable insights into the test’s sensitivity and specificity across a range of cognitive performances.

Finally, it may be worth considering conducting test-retest reliability analyses in future studies, although the literature suggests good long-term stability and the absence of a learning effect (Vyshedskiy et al., 2022) [7]. The ongoing longitudinal study of controls, MCI, and dementia will help determine long-term stability and the potential of BOCA as a tool for tracking cognitive changes over time.

## Conclusions

The study suggests that the Italian version of BOCA is a valid and applicable tool with a high discriminative ability for assessing cognitive status across the spectrum from normal cognition to dementia. The findings highlight the significance of early detection and monitoring of cognitive decline and suggest that BOCA could be a feasible and effective screening and monitoring tool in the population due to its stability and cost-efficiency. However, it is important to note that the study suggests that further extensive validation is necessary to explore the cost-effectiveness and applicability of BOCA as a cognitive assessment tool in aging individuals. This indicates the potential impact it may have on enhancing cognitive health assessments and care for aging populations.

## Data Availability

All data produced in the present work are contained in the manuscript

